# Pica in childhood: concurrent and sequential psychiatric comorbidity

**DOI:** 10.1101/2025.02.01.25321515

**Authors:** Laura Rubino, Cynthia Bulik, Samuel JRA Chawner, Nadia Micali

**Affiliations:** Department of Psychological and Brain Sciences, Drexel University; Centre for Neuropsychiatric Genetics & Genomics, School of Medicine, Cardiff University; Department of Psychiatry, University of North Caroline at Chapel Hill; Center for Eating and feeding Disorders research, Mental Health Services of the Capital Region of Denmark, Psychiatric Centre Ballerup, Denmark; Institute of Biological Psychiatry, Psykiatrisk Center Sct. Hans, Boserupvej 2, 4000 Roskilde, Denmark; Great Ormond Street Institute of Child Health, University College London, London, UK

**Keywords:** Pica, DAWBA, ADHD, eating disorders, comorbidity, ALSPAC

## Abstract

**Objective:** Pica is the persistent eating of non-nutritive, nonfood substances and is associated with serious medical consequences. There has been a lack of research into the psychiatric comorbidities of pica, despite being important for informing clinical care. The current study examines psychiatric comorbidities of pica in childhood and the longitudinal relationship between childhood pica and adolescent eating disorders. **Methods:** We analyzed data from the Avon Longitudinal Study of Parents and Children study. Pica and psychopathology, assessed with the Development and Well-Being Assessment and the Strengths and Difficulties Questionnaire, were assessed at about 7- and 10-years of age, reported eating disorders (ED) at 14-, 16-, and 18-years of age. We conducted linear and logistic regression models, adjusting for covariates, to identify concurrent psychiatric comorbidities, as well as risk for later EDs. **Results**. Pica was associated with increased odds of any psychiatric disorder and behavioral disorders in early childhood [OR = 7.30, *p* < .001 and OR = 5.65, *p* < .001, respectively] and mid-childhood [OR = 5.75, *p* < .001 and OR = 10.66, *p* < .001, respectively], and greater concurrent hyperactivity, conduct problems, peer problems, prosocial and emotional difficulties (*p* <.01 across analyses). We did not find evidence pica presence increased odds for concurrent emotional disorders nor for later ED risk. **Discussion**: The association between pica and psychiatric and behavioral disorders, indicates a likely shared aetiology. Our findings provide insight into the psychiatric characteristics of children with pica and highlight they may require complex behavioral support beyond their eating difficulties. [250]

**Public Significance (for IJED):** This study investigated the relationship between pica (the eating of non-food substances) and mental health conditions in childhood. We found that in childhood, pica was associated with increased odds of having behavioral disorders (e.g., ADHD and conduct disorder) and mental disorder (e.g., anxiety or ADHD) but not emotional disorders (e.g., OCD and depression). Our findings suggest children with pica may require complex behavioral support beyond their eating difficulties.

## Introduction

Pica was recognized as an official feeding and eating disorder in the fifth edition of the Diagnostic and Statistical Manual of Mental Disorders (DSM-5). It is characterized by the “persistent eating of non-nutritive, nonfood substances over a period of at least one month” (American Psychiatric Association (APA), 2013).” To be considered pica, the eating behavior must be inappropriate regarding the developmental level of the individual and not part of a culturally accepted practice. Further, if the eating behavior is comorbid with another psychiatric or medical condition (including pregnancy) it must be severe enough to garner additional clinical concern (DSM-5; APA, 2013). Pica is associated with serious medical side-effects, such as ruptures or obstructions in gastrointestinal systems, anemia, and increased mortality (Decker, 1993; Dumaguing et al., 2003; McLoughlin, 1988; Syrakos et al., 2008). Despite potentially serious consequences, pica remains understudied. It is essential we increase our understanding of pica and who might be more at risk of engaging in pica to inform identification and clinical management.

Studies investigating the prevalence of pica in children are heterogeneous regarding setting, sample characteristics, and diagnostic criteria, and report a range of prevalence estimates from 1.7% to 5.0% (Hartmann et al., 2018; Marchi & Cohen, 1990; Murray et al., 2018) and indicate pica prevalence might ameliorate as children grow older (Papini et al., 2024). Pica has been found to be more common in autistic children than non-autistic children throughout development (Emond et al., 2010; Papini et al., 2024), and pica behaviors have been reported to be present in 23% of autistic children (Fields et al., 2021). However it should be emphasized that pica does not exclusively occur in children with neurodevelopmental conditions, and is present in neurotypical children (Fields et al., 2021; Papini et al., 2024). The varying prevalence estimates may reflect true population differences, differences across developmental stages, but could also reflect methodological differences. To increase our understanding of who engages in or might be at risk for pica behavior, we need to better characterize the relationship between pica and other childhood psychopathology both concurrently and longitudinally (across development).

Although behaviors associated with pica have been linked to various behavioral and emotional disorders through case studies and hypothesized theoretical models, there is a lack of empirical large-scale studies investigating the association between pica and psychopathology. For example, pica has been conceptualized as a potential compulsive behavior (Bhatia & Gupta, 2009; Stein et al., 1996). Case studies found treatment for OCD led to a reduction in pica behavior (Bhatia & Gupta, 2009; Hergüner et al., 2008), but to our knowledge no study has investigated the prevalence of pica among children with OCD, nor whether children with pica are more likely to have OCD. Additionally, pica has also been conceptualized as being a disorder of impulse control (Saddichha et al., 2012). Case studies report medication treatment for ADHD decreased pica behavior (Hergüner & Hergüner, 2010; Saddichha et al., 2012; Vieira & Castello Branco, 2023). However in both autistic children (Neumeyer et al., 2019) and neurotypical children (Hartmann et al., 2018), pica was not significantly correlated to ADHD or hyperactivity, respectively. Furthermore, we are unaware of any studies directly investigating the relationship between pica and anxiety or depression in children.

Furthermore, there is mixed evidence that pica is cross-sectionally associated with other eating disorders and eating disorder behaviors. Although one study reported pica was not associated with dietary restraint and weight and shape concerns in children (Hartmann et al., 2018), others studies found pica was associated with greater fears of weight gain, dietary restriction, dissatisfaction with shape and weight, purging, and binge eating in children (Murray et al., 2018) and adults (Hartmann et al., 2022). Within AN, some individuals eat ice and non-food items, but whether pica precedes or antecedes development of AN is unclear (DSM-5). Work is needed to confirm the relationship between pica and whether pica behavior in childhood is associated with later ED diagnoses (i.e. sequential comorbidity).

The current study aims to investigate the overlap between pica and other psychiatric conditions. The second aim is to examine the prospective association between pica in childhood and eating disorders diagnoses in mid to late adolescence.

## Methods

### Participants

We accessed data from the Avon Longitudinal Study of Parents and Children (ALSPAC), a cohort-based longitudinal study that invited all pregnant women expected to give birth during April 1, 1991 and December 31, 1992, living in or near Avon, UK to participate. 14,541 children from these pregnancies were enrolled; 13,988 children were alive at 1 year. At the 7-year assessment, 713 more children were enrolled. At the late childhood (after 7-year) assessment, 151 additional children were enrolled, for a total of 15,447 pregnancies included after age 7. Data were available from 13,906 children at the 7 year assessment, 12,772 children in late childhood (> 7 and <13 years old), 12,254 during adolescents (13-16 years old), and 11,875 children from the transition to adulthood period (16 to 18 years old; Boyd et al., 2013; Fraser et al., 2013). The total sample size for analyses using any data collected after the age of seven is therefore 15,447 pregnancies, resulting in 15,658 foetuses. Of these 14,901 children were alive at 1 year of age. All women gave informed consent. Informed consent for the use of data collected via questionnaires and clinics was obtained from participants following the recommendations of the ALSPAC Ethics and Law Committee at the time. Teenagers were re-consented when they reached 18 years of age. If consent was withdrawn at any time point, the participant’s data was not used from that time point on. Ethical approval for the study was obtained from the ALSPAC Ethics and Law Committee and the Local Research Ethics Committees. For this study, we excluded children for which sex, i.e. sex assigned at birth, data was not available. Further, we removed one twin from twin pairs at random (Micali et al., 2015).

### Procedure

Mothers were recruited through media advertisements and conversations at community locations and with doctors at maternity health services. Once enrolled and consented, there were frequent assessments between the child’s birth and them turning 18 years of age. Assessments included child-completed questionnaires, clinical assessments, and assessments about the child completed by the mother or other main caregiver (Boyd et al., 2013). The current study uses questionnaires the mother or main caregiver completed about pica behaviors in their child at the 36-, 54-, 65-, 77-, and 115-month assessments and psychiatric disorders in their child at the 7-year (81 and 91 months) and 10-year (108 and 128 months) assessments; and eating disorder diagnoses were obtained from child, parent, and objective data at the 14-year, 16-year, and 18 year wave (Micali et al., 2015). Please note that the study website contains details of all the data that is available through a fully searchable data dictionary and variable search tool: http://www.bristol.ac.uk/alspac/researchers/our-data/.

### Measures

#### Pica

At 36-, 54-, 65-, 77-, and 115-month mothers were asked, “How often does the child eat coal, dirt, or other non-nutritious substances?” Responses options were rated on a scale of one to four, representing “yes, every day,” “yes, at least once a week,” “yes, less than once a week,” and “no, not at all,” respectively. If respondents reported at least “yes, less than once a week,” they were asked a follow-up question to clarify what the child ate to ensure understanding of the question. Following previous work, we created a binary pica yes/no value at each time point (see Papini et al., 2024 for details). We also created a variable indicating persistent pica (pica_pers:_ if pica was present at two timepoints or more across the 5 assessments) and any pica (pica_any:_ if pica was present at least at one timepoint).

#### Psychiatric Disorders

At the 91- and 128-months (child’s age), parents completed the Development and Well-Being Assessment (DAWBA; Goodman et al., 2000) answering questions about their child, to generate International Classification of Diseases-10 (ICD-10) and Diagnostic and Statistical Manual of Mental Illness, fourth edition (DSM-IV) psychiatric diagnoses. For this study, the DAWBA was adapted to be a questionnaire instead of a structured interview designed to generate the likelihood of the presence of psychiatric illnesses (University of Bristol). Following an established and validated approach, a computerized algorithm was applied to the caregiver responses, deriving the likelihood of a diagnosis, scored on a six-point scale based (0 = 0.1%; 1 = ∼ 0.5%; 2 = ∼ 3%, 3 = 15%, 4 = 50%, 5 = ≥ 70%; (Goodman et al., 2000; Youthinmind, 2022). From these likelihoods, a binary yes/no presence of disorder was derived.

#### Dimensional Psychopathology

At the 81- and 108-month child age, mothers or main caregivers completed the strengths and difficulties questionnaire (SDQ; Goodman, 1997). The total scores for the emotional symptoms, conduct problems, hyperactivity/inattention, peer relationship problems subscales range from zero to ten with higher scores indicating greater psychological difficulties. The prosocial behavior subscale is also scored on a scale of zero to ten, but higher scores indicate lower psychological difficulties. A total difficulties score consisting of all subscales but prosocial was also computed, scores range from 0 to 40 with higher scores indicating greater difficulties.

#### Eating Disorder Symptoms

At the 14-, 16-, and 18-years youth completed questionnaires assessing ED behaviors. Micali et al., 2015 derived eating disorder diagnoses for the adolescents using these data, parental data and objective weight and height (see Micali et al., 2015).

Adolescents were categorized as having no eating disorder, full-threshold Binge Eating Disorder (BED), Sub-threshold (S-BED), full-threshold Bulimia Nervosa (BN), sub-threshold BN (S-BN), anorexia nervosa (AN), and purging disorder (PD; Micali et al., 2015). We also created any “any age”

### Statistical Analyses

#### Descriptive analysis

Prevalence of pica at the at 36-, 54-, 65-, 77-, and 115-month assessments (Papini et al., 2024) and EDs at 14- and 16-year assessments (Micali et al., 2015) in the current sample have been previously reported. We calculated the frequency of EDs at 18 years of age based on definitions used in Micali et al., 2015 for EDs at 14 and 16 years of age. We examined the frequency of various DAWBA diagnoses and created cross-tabulation tables for pica at 77- and 115-months and DAWBA diagnoses (OCD, any anxiety disorder, any depressive disorder, any ADHD disorder, oppositional conduct disorder, pervasive developmental disorder) at 91- and 128-months, respectively. We also created cross-tabulation tables for ED diagnoses at the 14-, 16-, and 18-year assessments and any pica and persistent pica.

After examining cross-tabulations tables between pica and the separate DAWBA diagnoses, we observed very low cell counts not appropriate for further analyses. To protect participant confidentiality, and in alignment with the ALSPAC protocols, we have not reported these tables. Given low cell counts, we combined DAWBA diagnoses into three categories: any behavioral, any emotional, and any disorder at both 91- and 28-months of age. Any behavioral consisted of any ADHD, oppositional conduct, pervasive developmental disorder. Any emotional consisted of OCD, any anxiety disorder, and any depressive disorder. Any Disorder included all disorders in both any behavioral and any emotional. We used these combined categories instead of individual disorder diagnoses in subsequent analyses.

We also observed low cell counts between pica_pers_ and ED outcomes. Consequently, we did not conduct logistic regressions using pica_pers_ as the predictor, and only examined the prospective relationship between pica_any_ and EDs. Further, we collapsed ED diagnostic categories into full threshold, full and subthreshold, and any ED. Full threshold includes those with full-threshold BED, BN, AN, or PD. Full and subthreshold includes those with full-threshold BED, BN, AN, or PD, and sub-threshold BED and BN. Any ED includes those with full-threshold BED, BN, AN, or PD, and sub-threshold BED and BN, and those with EDNOS. All categories are as defined by Micali et al., 2015. We also collapsed timepoints, and “any age,” such that if an individual had full-threshold, sub-threshold, or any ED at any age it was coded present, and if not, was coded absent.

Table 1 presents the cross-tabulation tables for behavioral, emotional, any DAWBA diagnosis, and EDs with pica across assessments.

**Table 1.** Frequency of co-occurring conditions, stratified by, pica presence.

| DAWBA Diagnoses |  |  | <u>Any behavioral</u> |  | <u>Any emotional</u> |  | <u>Any DAWBA</u> |  |
| --- | --- | --- | --- | --- | --- | --- | --- | --- |
|  |  |  | Absent | Present | Absent | Present | Absent | Present |
| 77 months | absent |  | 6868 | 240 | 6999 | 132 | 6756 | 375 |
|  | present |  | 34 | **7** | 38 | **<5\*** | 29 | **12** |
| 115 months | absent |  | 6330 | 210 | 6411 | 177 | 6217 | 317 |
|  | present |  | 15 | **6** | 21 | **<5\*** | 16 | **6** |
|  |  |  | <u>Full threshold<sup>1</sup></u> |  | <u>Full and Sub threshold<sup>2</sup></u> |  | <u>Any ED psychopathology<sup>3</sup></u> |  |
| Eating Disorders |  |  | Absent | Present | Absent | Present | Absent | Present |
| Age 14 | Persistent | Absent | 4053 | 162 | 4005 | 210 | 2490 | 725 |
|  | Pica<sup>4</sup> | Present | 24 | **<5\*** | 24 | **<5\*** | 21 | **<5\*** |
|  | Any Pica | Absent | 3964 | 159 | 3917 | 206 | 3421 | 702 |
|  |  | Present | 113 | **<5** | 112 | **<5\*** | 90 | **26** |
| Age 16 | Persistent | Absent | 3408 | 184 | 3291 | 301 | 2299 | 1293 |
|  | Pica<sup>4</sup> | Present | 18 | **<5\*** | 15 | **<5\*** | 7 | **11** |
|  | Any Pica | Absent | 3325 | 180 | 3210 | 295 | 2239 | 1266 |
|  |  | Present | 101 | **<5\*** | 96 | **>5\*** | 67 | **38** |
| Age 18 | Persistent | Absent | 2300 | 124 | 2177 | 247 | 2133 | 291 |
|  | Pica | Present | 16 | **<5\*** | 15 | **<5\*** | 15 | **<5\*** |
|  | Any Pica | Absent | 2238 | 123 | 2120 | 241 | 2079 | 282 |
|  |  | Present | 78 | **<5\*** | 72 | **>5\*** | 69 | **10** |
| Any age<sup>5</sup> | Persistent | Absent | 1773 | 403 | 1637 | 627 | 1079 | 1794 |
|  | Pica | Present | 11 | **<5\*** | 9 | **<5\*** | 5 | **12** |
|  | Any Pica | Absent | 1726 | 395 | 1595 | 612 | 1048 | 1750 |
|  |  | Present | 58 | **8** | 51 | **19** | 36 | **56** |

\*Cell counts <5 may include zero
<sup>1</sup> Full threshold includes those with full-threshold BED, BN, AN, or PD, defined by Micali et al., 2015<sup>2</sup> Full and subthreshold includes those with full-threshold BED, BN, AN, or PD, and sub-threshold BED and BN, defined by Micali et al., 2015<sup>3</sup> includes those with full-threshold BED, BN, AN, or PD, and sub-threshold BED and BN, and those with EDNOS and at-risk for EDs, defined by Micali et al., 2015<sup>4</sup> Participants were coded to have any pica if they endorsed pica behaviors at any timepoint in childhood. Persistent pica represents pica behaviors endorsed at at least 2 time points<sup>5</sup> Any age is collapsed across ages 14, 16, or 18. For example, any age full threshold is individuals who had full threshold at 14, 16, and/or 18.

#### Aim 1

We conducted six logistic regression models to assess the relationship between pica and DAWBA diagnoses. We conducted three models for early childhood. Each used binary pica presence at 77 months for the predictor and a different DAWBA diagnosis (behavioral disorders, any emotional, and any DAWBA) at 91 months for the outcome. We conducted three models for mid-childhood. Each mid-childhood model used binary pica presence at 115 months for the predictor and a different DAWBA diagnosis (behavioral disorders, any emotional, and any DAWBA) at 91 months for the outcome. All models included sex as a covariate.

We conducted 12 linear regression models to assess the relationship between pica and SDQ scores. Six models were run for early-childhood. Each early childhood model used binary pica presence at 77 months as the predictor, and a different SDQ subscale score at or the SDQ total score at 81 months for the outcome. Six models were run for mid-childhood. Each early childhood model used binary pica presence at 115 months as the predictor, and a different SDQ subscale score at or the SDQ total score at 108 months for the outcome. All 12 models included sex as a covariate. Before conducting the linear regressions we tested the assumptions of linear regression and found the normality of residuals assumption was violated and that subscale scores exhibited skewness and kurtosis values outside of the acceptable range. To address these issues, we Box-Cox transformed (using MASS R package; Venables & Ripley, 2002) all the SDQ subscales at each time-point and standardized to z-scores. The z-scores were then included as the outcome variable within the linear regression models. We calculated *sr^2^*, the squared semi-partial correlation, representing variance explained by that unique predictor in the model for each analysis.

#### Aim 2

We conducted 12 logistic regressions to assess the relationship between pica (pica_any_,) in childhood and EDs later in adolescence. There were four models for full threshold ED, four models for full threshold + sub threshold, and four models for any ED. Each model used pica_any_ as the predictor, and full threshold, full and subthreshold, and any ED at either 14, 16, 18 years of age, or any age as the outcome. All analyses controlled for sex.

#### Post hoc-exploratory analyses

After conducting the analyses for Aims 1 and 2, we conducted exploratory analyses to control for potential effects of child developmental level. At 42 months of age, children were assessed with the Denver Developmental Screening Test (Frankenburg & Dodds, 1967). This screening tool helps identify children with developmental delays in personal-social, fine motor and adaptive, language, and gross motor domains. We categorized the lowest decile as indicative of developmental delay (Papini et al., 2024). We then added this binary variable as a covariate in the Aim 1 SDQ Analyses.

All tests were conducted in R using the lmer package. We used listwise deletion, such that for each separate analysis we only included people who completed both measures. Therefore, each analysis varies in missingness.

## Results

### Descriptives

The prevalence of pica at the various ages was 2.33% at 38 months, 0.79% at 54 months, 0.65% at 65 months, 0.56% at 77 months, 0.33% at 115 months. 3.10% of participants experienced pica_any_, and 0.65% had pica_pers_.

Supplemental Table 3 presents the frequencies of DAWBA diagnoses at the 91- and 120-month assessments, and EDs at 14-, 16-, and 18-years of age assessments.

#### Psychopathology

In early childhood, individuals with pica had greater odds of having any behavioral disorder [OR = 5.65, 95% CI = 2.23-12.43, *p* < .001] and greater odds of having any psychiatric disorder [OR = 7.30, 95% CI = 3.53-14.22, *p* < .001] than individuals without pica. In early childhood, individuals with pica also had greater odds of having any emotional disorder, despite a wide CI [OR = 4.07, 95% CI = 0.97-11.45, *p* < .006]. A CI including zero typically indicates non-significance. The discrepancy of the CI including zero but a *p* <.05 could be due to the low number of cases of individuals with both pica and any emotional disorder (Table 2), so this result should be treated cautiously.

**Table 2.** Logistic Regression Results.

| | | B | SE | p | OR | 95% CI of OR | $\chi^2$ | Wald's test | AIC | Df model |
| --- | --- | --- | --- | --- | --- | --- | --- | --- | --- | --- |
| 77 months | Any behavioral | 1.73 | 0.43 | <.001 | 5.65 | [2.23, 12.43] | 91.4 | 0.0 | 2046.6 | 7135 |
|  | Any emotional | 1.40 | 0.61 | 0.02 | 4.07 | [0.97, 11.45] | 10.3 | .006 | 1336.9 | 7158 |
|  | Any DAWBA | 1.99 | 0.35 | <.001 | 7.30 | [3.53, 14.22] | 79.6 | <.001 | 2936 | 7158 |
| 115 months | Any behavioral | 2.37 | 0.49 | <.001 | 10.66 | [3.73, 26.86] | 54.6 | <.001 | 1848.8 | 6550 |
|  | Any emotional | 0.54 | 1.02 | .60 | 1.71 | [0.10, 8.27] | 0.31 | .86 | 1643.2 | 6599 |
|  | Any DAWBA | 1.75 | 0.48 | <.001 | 5.75 | [2.05, 14.15] | 32.3 | <.001 | 2862.1 | 6599 |

In mid-childhood, individuals with pica, had greater odds of having any behavioral disorder [OR = 10.66, 95% CI = 3.73, 26.86, *p* < .001], and greater odds of having any psychiatric disorder [OR = 5.75, 95% CI = 2.05, 14.15, *p* < .001] than individuals without pica. In mid-childhood, individuals with pica also had greater odds of having any emotional disorder, although this difference was not statistically significant [OR = 1.71 95% CI = 0.10-8.27, *p* =.86], (Table 2).

#### Dimensional Psychopathology

In early childhood, pica was significantly associated with all SDQ subscales and total SDQ score (Table 3). *R^2^* values were negligible for the emotional [*b* = 0.389, *sr^2^=*.00, *R^2^*=0.004, *p*<.01], conduct [*b* = 0.88, *sr^2^=*.00, *R^2^*=0.009, *p*<.01], and peer problems [*b* = 0.74, *sr^2^=*.00, *R^2^*=0.007, *p*<.01] subscales and small for the prosocial [*b* = −0.80, *sr^2^=*.00, *R^2^*=0.036, *p*<.01], hyperactivity [*b* = 0.83, *sr^2^=*.00, *R^2^*=0.031, *p*<.01], and total difficulties [*b* = 1.03, *sr^2^=*.01, *R^2^*=0.017, *p*<.01] subscales (Table 3; Cohen, 1988). The presence of Pica corresponded to a 0.88, 0.39, 0.83, 0.74, and 1.03 z-score increase on the conduct problems, emotional problems, hyperactivity, peer problems, and total difficulties subscales respectively, compared to no pica. Pica also corresponded to 0.80 z-score points lower on the prosocial scale.

**Table 3.** Linear Regression Results, SDQ subscales as the outcome.

|  |  | 77 months |  |  |  |  | 115 months |  |  |  |  |
| --- | --- | --- | --- | --- | --- | --- | --- | --- | --- | --- | --- |
|  | Predictor | *b* | *beta* | *sr*<sup>2</sup> | *r* | Fit | *b* | *beta* | *sr*<sup>2</sup> | *r* | Fit |
| Conduct Problems | Pica | 0.88** | 0.06 | .00 | .07** |  | 0.76** | 0.04 | .00 | .05** |  |
|  | sex | -0.13** | -0.07 | .00 | -.07** |  | -0.10** | -0.05 | .00 | -.05** |  |
| | Model fit | | | | | $R^2 = .009^{**}$<br>95% CI[.00,.01] | | | | | $R^2 = .005^{**}$<br>95% CI[.00,.01] |
| Emotional Problems | Pica | 0.39* | 0.03 | .00 | .03* |  | 0.95** | 0.05 | .00 | .05** |  |
|  | sex | 0.11** | 0.05 | .00 | .05** |  | 0.17** | 0.08 | .01 | .08** |  |
| | Model fit | | | | | $R^2 = .004^{**}$<br>95% CI[.00,.01] | | | | | $R^2 = .010^{**}$<br>95% CI[.00,.01] |
| Hyperactivity | Pica | 0.83** | 0.06 | .00 | .06** |  | 1.23** | 0.07 | .01 | .08** |  |
|  | sex | -0.33** | -0.16 | .03 | -.17** |  | -0.33** | -0.17 | .03 | -.17** |  |
| | Model fit | | | | | $R^2 = .031^{**}$<br>95% CI[.02,.04] | | | | | $R^2 = .033^{**}$<br>95% CI[.03,.04] |
| Peer problems | Pica | 0.74** | 0.05 | .00 | .05** |  | 0.94** | 0.05 | .00 | .06** |  |
|  | sex | -0.12** | -0.06 | .00 | -.06** |  | -0.07** | -0.04 | .00 | -.04** |  |
| | Model fit | | | | | $R^2 = .007^{**}$<br>95% CI[.00,.01] | | | | | $R^2 = .004^{**}$<br>95% CI[.00,.01] |
| Prosocial | Pica | -0.80** | -0.06 | .00 | -.06** |  | -0.44* | -0.03 | .00 | -.03** |  |
|  | sex | 0.36** | 0.18 | .03 | .18** |  | 0.39** | 0.19 | .04 | .19** |  |
| | Model fit | | | | | $R^2 = .036^{**}$<br>95% CI[.03,.04] | | | | | $R^2 = .038^{**}$<br>95% CI[.03,.05] |
| Total Difficulties | Pica | 1.03** | 0.07 | .01 | .08** |  | 1.45** | 0.08 | .01 | .09** |  |
|  | sex | -0.22** | -0.11 | .01 | -.11** |  | -0.15** | -0.08 | .01 | -.08** |  |
| | Model fit | | | | | $R^2 = .017^{**}$<br>95% CI[.01,.02] | | | | | $R^2 = .013^{**}$<br>95% CI[.01,.02] |

\* $p < .05$ , \*\* $p < .01$ , \*\*\* $p < .001$ **Table 4.** Logistic Regression Results, Predictor: Any Pica<sup>1</sup>, Outcome: ED

In mid-childhood, pica was significantly associated with each SDQ subscale examined and total SDQ score. *R^2^* values were negligible for the emotional [*b* = 0.95, *sr^2^=*.00, *R^2^*=0.010, *p*<.01], conduct [*b* = 0.76, *sr^2^=*.00, *R^2^*=0.005, *p*<.01], and peer problems [*b* = 0.94, *sr^2^=*.00, *R^2^*=0.004, *p*<.01] subscales, and small for the prosocial [*b* = −0.44, *sr^2^=*.00, *R^2^*=0.038, *p*<.01] and hyperactivity subscales [*b* = 1.23, *sr^2^=*.00, *R^2^*=0.033, *p*<.01] and total difficulties score [*b* = 1.45, *sr^2^=*.01, *R^2^*=0.013, *p*<.01] (Table 3). Results indicate that Pica presence corresponded to 1.19, 1.19, 1.23, 0.94, and 1.45 z-score points higher on the conduct problems, emotional problems, hyperactivity, peer problems, and total difficulties subscales respectively, indicating greater problems for children with pica. Pica also corresponded to 0.44 z-score points lower on the prosocial scale, indicating worse prosocial skills for children with pica.

For both time periods, although the z-score differences between groups were moderate to large between the groups, both *R^2^* and *sr^2^* values associated with pica were negligible to small, indicating that pica explains a negligible to small amount of the variance in SDQ scores within the ALSPAC population (0.00 to 0.01; Table 3) and other variables contribute to SDQ score outcomes.

### Aim 2

Logistic regression analysis revealed that the presence of pica did not significantly increase odds of having a full threshold, sub threshold, or any ED diagnosis at 14-, 16-, and 18-years of age (Table 4).

**Table 4.** Logistic Regression Results, Predictor: Any Pica^1^, Outcome: ED.

|  |  | B | SE | p | OR | AIC | Df model |
| --- | --- | --- | --- | --- | --- | --- | --- |
| Full threshold<sup>2</sup> | Age 14 | -0.41 | 0.59 | .49 | 0.66 | 1359.2 | 4236 |
|  | Age 16 | -0.36 | 0.52 | .49 | 0.70 | 1392.6 | 3607 |
|  | Age 18 | -1.49 | 1.01 | .14 | 0.22 | 947.88 | 2437 |
|  | Any age | -0.49 | 0.38 | .20 | 0.61 | 2043.3 | 2184 |
| Full + sub threshold<sup>3</sup> | Age 14 | -0.38 | 0.52 | .46 | 0.68 | 1648.5 | 4236 |
|  | Age 16 | -0.02 | 0.36 | .96 | 0.98 | 1981.7 | 3607 |
|  | Age 18 | -0.18 | 0.41 | .65 | 0.83 | 1556.6 | 2437 |
|  | Any age | -0.02 | 0.38 | .94 | 0.98 | 2615.9 | 2274 |
| Any ED diagnosis<sup>4</sup> | Age 14 | 0.36 | 0.23 | 0.12 | 1.43 | 3801.2 | 4236 |
|  | Age 16 | -0.05 | 0.22 | 0.84 | 0.96 | 434.9 | 3607 |
|  | Age 18 | 0.04 | 0.35 | 0.91 | 1.04 | 1723 | 2437 |
|  | Any age | -0.09 | 0.22 | 0.69 | 0.91 | 3668.4 | 2887 |

<sup>1</sup>Participants were coded to have any pica if they endorsed pica behaviors at any timepoint in childhood
<sup>2</sup>Full threshold includes those with full-threshold BED, BN, AN, or PD, defined by Micali et al., 2015
<sup>3</sup>Full and subthreshold includes those with full-threshold BED, BN, AN, or PD, and sub-threshold BED and BN, defined by Micali et al., 2015
<sup>4</sup>includes those with full-threshold BED, BN, AN, or PD, and sub-threshold BED and BN, and those with EDNOS, defined by Micali et al., 2015

### Post hoc analyses

Given the large mean differences observed between those with and without pica on the SDQ subscales, but low variation explained in the linear regressions, we investigated whether having a developmental disability explained some of the difference found, by including developmental disability as a covariate. See Table 5 for results.

**Table 5.**
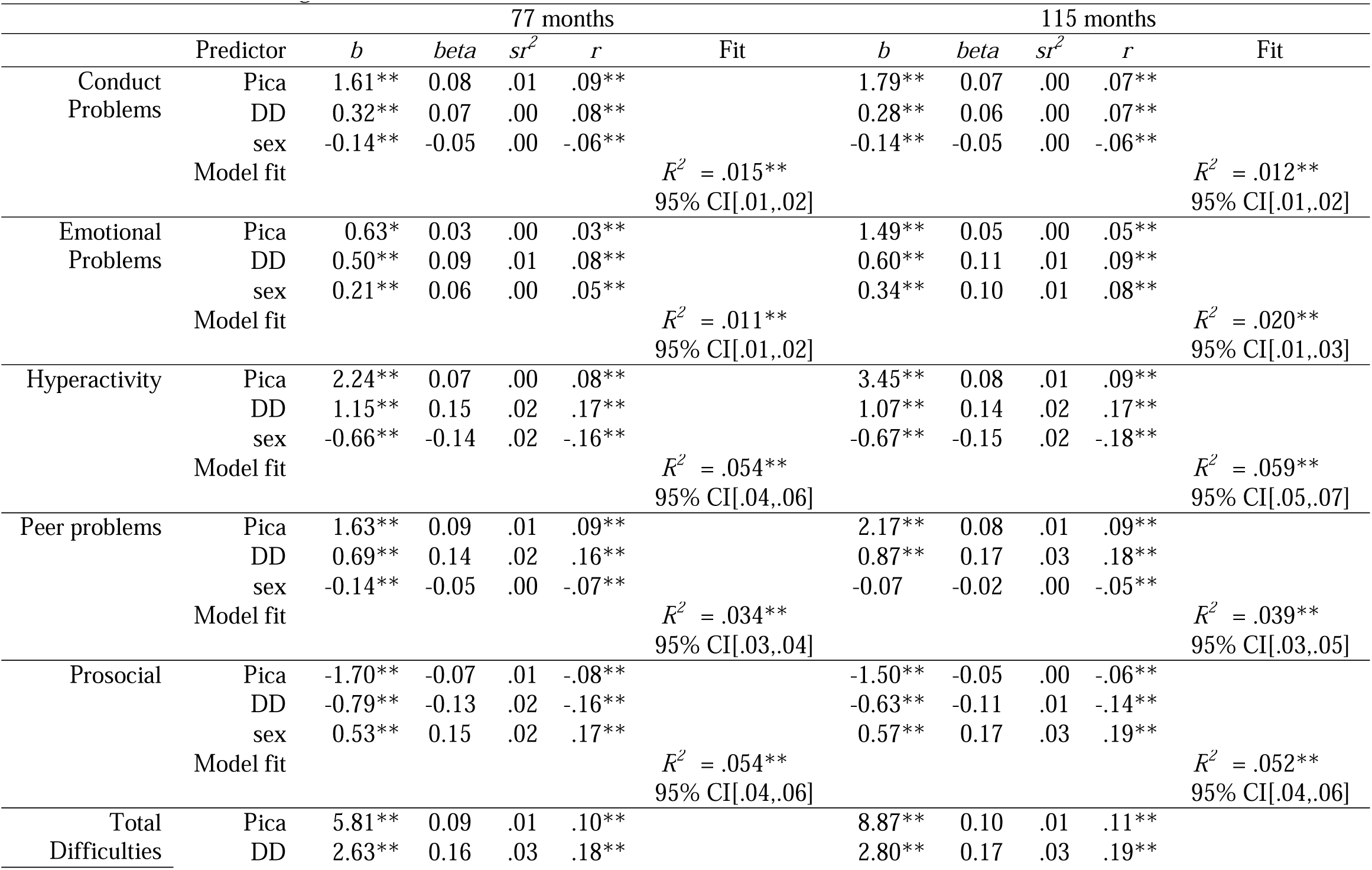
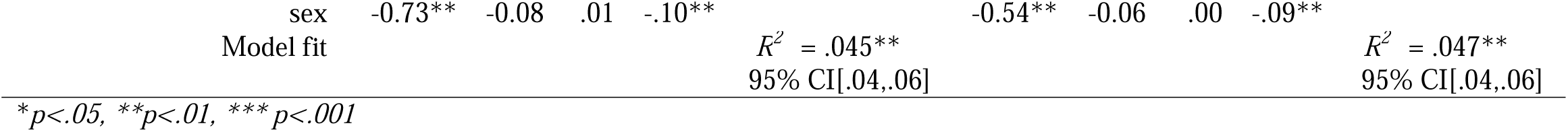
Post Hoc Linear Regression Results, SDQ subscales as the outcome.

Pica remained a significant predictor in the model after taking developmental delay into account. The R^2^ values for each regression model did increase with the addition of developmental delay as a predictor. However, the R^2^ values remained negligible to small, indicating the model explained a negligible to small percent (ranging from 1.2% to 5.4%), of the difference in means scores observed between those with and without pica. Further, the partial variance explained by the presence of a developmental disability was still negligible to small, with *sr^2^* ranging from 0.00 to 0.03 (see Table 5).

## Discussion

The current study aimed to increase our understanding of psychiatric comorbidities of pica in childhood and later risk of eating disorder onset if pica was present in childhood. This is the first study to report concurrent comorbidities at more than one timepoint throughout development and the association between pica and later eating disorders. Furthermore, by using the ALSPAC dataset, a large-scale population cohort, the current study provides improved estimates of psychiatric comorbidities of pica compared to potential biases potentially present in previous heterogeneous small-scale studies. We found caregiver endorsement of pica behaviors in childhood (in early and mid-childhood) increased odds of having a concurrent behavioral problem (ORs = 5.65 and 7.30, respectively) or any psychiatric problem (ORs = 10.66 and 5.75, respectively) at the diagnostic level (as measured by the DAWBA). We also found children with pica had greater conduct, emotional, prosocial, hyperactivity, and peer problems than children without pica. However, presence of pica behaviors did not significantly increase the odds of having an emotional disorder diagnosis. Furthermore, our findings suggest engaging in pica behaviors at any point throughout childhood was not associated with increased odds of developing an eating disorder, specifically BED, BN, AN, or PD in adolescence (14, 16, or 18 years of age).

Findings from the current study suggests pica may be associated with risk of behavioral disorders rather than emotional disorders, indicating there might be common aetiological mechanisms between pica and behavioral disorders. Pica did not statistically significantly increase odds of an emotional disorder at the diagnostic level but there was a small difference (z-score mean difference of 0.39 in early childhood and 1.23 in mid-childhood) in subthreshold dimensional psychopathology. The association between pica and emotional traits is significant but small (see aforementioned effect size). This indicates pica may be associated with emotional psychopathology at a trait level, but the effect size is not large enough for an individual to meet diagnostic criteria for an emotional disorder.

In these analyses, we had to collapse categories, meaning we were unable to investigate specific mental health disorders. However, in the current study, pica corresponded to the greatest mean difference in hyperactivity out of the behavioral subscales measured by the SDQ. This analysis of dimensional traits provides a fine grain investigation in addition to the diagnostic-level analyses. These findings supports the theory that pica may be associated with greater ADHD and impulsivity, as children with pica in the current study reported greater hyperactivity—a key trait of ADHD—than children without pica (Hergüner & Hergüner, 2010). The current study shows stronger support for a relationship between hyperactivity and pica rather than compulsivity and pica, as cross-tabulation cells counts were low between pica and OCD and did not permit logistic regression analyses potentially indicating low-comorbidity. Further, pica presence remained a significant predictor of psychopathology, even after we added developmental delay which, although significant, accounted for a small proportion of the variance. This study provides preliminary evidence that pica may be related to behavioral disorders and children with pica may struggle with more behavioral disorders.

The current results also suggest pica has broad effects on psychopathology including peer relations, pro-sociality, emotional, and behavioral health. Although pica and developmental delay explained only negligible to small amounts of the variance observed in behavioral problems between those with pica and those without pica, the moderate to large differences in mean scores imply although individuals with pica are at increased risk of behavioral problems. This indicates that screening for behavioral problems within individuals with pica is more likely to be an effective screening strategy than screening pica within children with behavioral problems.

Results also suggest individuals with pica are not at increased odds of developing BED, BN, PD, or AN in mid to late adolescence. Unfortunately, the ALSPAC dataset did not contain information on ARFID and rumination diagnosis. Thus, we were unable to assess the association between pica and adolescent ARFID and rumination, which have previously shown the greatest overlap with pica (Hartmann et al., 2018; Murray et al., 2018). ARFID and pica are understood as feeding disorders, whereas BED, BN, PD, and AN are considered eating disorders in the DSM-5. Therefore, it may be that feeding disorders could overlap with each other more than feeding and eating disorders with each other. Results from other studies support the hypothesis that ARFID and pica share more overlap than with other eating disorders. Although our data suggest that there is not an increased risk of developing an eating disorder like AN, BN, PD, or BED, it will be important to replicate this finding in other population cohorts.

The study did have some limitations. First, although we had items to report pica behaviors, we acknowledge limitations in our measurement. As discussed in Papini et al., 2024, we could not derive DSM-5 Pica diagnosis. Further, the self-report items assessing pica limited the precision in which we could assess the frequency and severity of pica behaviors. Second, although the ALSPAC sample is large, we experienced small cell counts of symptom presentation and overlap which limited statistical analysis options and statistical power. The small cell counts also limited the detail and nuance to which we could investigate psychiatric comorbidities. However, this was, in part, mitigated by the inclusion of dimensional psychopathology measures. We also could not assess true concurrent existence of pica and psychopathology as the constructs were measured a few months apart in ALSPAC. Additionally, as noted in Papini et al., 2024, the ALSPAC cohort differs from the general population in the UK in that it is less diverse across socioeconomic status, race, and ethnicity than the general UK, potentially limiting generalizability of the sample. It is also possible the prevalence and characteristics of pica have shifted since the start of the ALSPAC study, and children who present with pica exhibit different characteristics than children with pica born in the early twentieth-first century.

To further our understanding of pica, future work would benefit from the creation of a questionnaire designed specifically to assess the diagnostic criteria of pica as described in the DSM-5, potentially adapted from such measures such as the Pica, ARFID, and Rumination Disorder Interview (PARDI; Bryant-Waugh et al., 2022). Most questionnaires, as with the current study, assess for the consumption of non-food substances but do not assess the other DSM-5 diagnostic criteria of pica, including duration (i.e., at least 1 month) and inappropriateness given developmental age. Given the low prevalence of pica, future work would benefit from Likert-type scales assessing frequency and severity of pica behaviors to increase statistical power and allow for a more precise understanding of pica behavior engagement and related characteristics. Further, future work should look at the longitudinal relationship between ARFID and pica and other eating disorders (e.g., Binge Eating Disorder and Bulimia Nervosa), across development.

Overall, the current study provides preliminary evidence of the association of pica with behavioral disorder diagnoses and a broad range of childhood psychopathology. This furthers our knowledge of the clinical presentation of this understudied feeding & eating disorder, and highlights the complex needs of individuals living with pica and indicates the need for psychiatric screening when pica presents clinically. This initial work provides the basis for future work investigating psychiatric comorbidities in pica, but also to understand the potential aetiological overlap of pica and behavioral disorders.

## Data Availability

Data is available given approval of a third party, ALSPAC.

## Abbreviations

ADHD: attention deficit hyperactivity disorder
ALSPAC: Avon Longitudinal Study of Parents and Children
AN: anorexia nervosa
BED: binge eating disorder
BN: bulimia nervosa
DSM-5: Diagnostic and Statistical Manual of Mental Disorders-Fifth Edition
ED: Eating Disorder
PD: purging disorder

## Acknowledgments

We are extremely grateful to all the families who took part in this study, the midwives for their help in recruiting them, and the whole ALSPAC team, which includes interviewers, computer and laboratory technicians, clerical workers, research scientists, volunteers, managers, receptionists and nurses. This work was funded by the Medical Research Foundation (MRF-058-0015-F-CHAW). We are also extremely grateful to the Center for Excellence in Eating Disorder (CEED) Summer Fellowship at the University of North Carolina at Chapel Hill for providing the funding and opportunity to collaborate for this study.

