## SupplementalTables for "Pica in childhood: concurrent and sequential psychiatric comorbidity"

| ***Table 1.*** *Prevalence of pica across development* | | |
| --- | --- | --- |
| Timepoint | Count (n) | Frequency |
| 38 months | 230 | 2.33% |
| 54 months | 75 | 0.79% |
| 65 months | 56 | 0.64% |
| 77 months | 47 | 0.56% |
| 115 months | 25 | 0.33% |
| Persistant^1^ | 39 | 0.65% |
| Any^2^ | 190 | 3.10% |
| ^1^P*ersistant pica was coded as present if pica was endorsed at two or more time points*  *^2^Any pica was coded as present if pica was endorsed at any time point* | | |

| ***Table 2.*** *DAWBA^1^ diagnosis frequency* | | | | |
| --- | --- | --- | --- | --- |
|  | 7 years old | | 10 years old | |
|  | Count | Proportion | Count | Proportion |
| OCD^2^ | 9 | 0.11% | <5 | 0.05% |
| Any anxiety | 251 | 3.10% | 152 | 2.07% |
| Any depressive | 39 | 0.48% | 70 | 0.97% |
| Any emotional | 173 | 2.14% | 202 | 2.74% |
| Any ADHD^2^ | 172 | 2.13% | 118 | 1.61% |
| Any Disorder | 463 | 5.85% | 437 | 6.02% |
| *^1^DAWBA = Development and Well-Being Assessment (Goodman et al., 2000)*  *^2^OCD = obsessive compulsive disorder*  *^3^ADHD = attention deficit/hyperactivity disorder* | | | | |

| ***Table 3.*** *Prevalence of eating disorders* | | | | | |  |  |  |  |
| --- | --- | --- | --- | --- | --- | --- | --- | --- | --- |
|  |  | Age 14 | | Age 16 | | Age 18 | | Any Age*^4^* | |
|  |  | Count (*n*) | Percent | Count (*n*) | Percent | Count | Percent | Count | Percent |
| Full threshold^1^ | BED | 28 | 0.48 | 55 | 1.14 | 53 | 1.67 | 121 | 4.80 |
|  | BN | 16 | 0.27 | 38 | 0.79 | 24 | 0.75 | 197 | 7.67 |
|  | PD | 24 | 0.41 | 75 | 1.55 | 54 | 1.70 | 137 | 5.40 |
|  | AN | 147 | 2.51 | 88 | 1.82 | 42 | 1.32 | 241 | 9.35 |
| Subthreshold^2^ | BED | <5* | NA | 23 | 0.48 | 140 | 4.40 | 163 | 6.49 |
|  | BN | 77 | 1.31 | 160 | 3.31 | 21 | 0.66 | 117 | 4.66 |
| Any ED^3^ |  | 1595 | 27.24 | 1929 | 39.90 | 1248 | 39.22 | 3277 | 76.89 |
| **cell counts <5 may include zero*  *^1^ Full threshold includes those with full-threshold BED, BN, AN, or PD, defined by Micali et al., 2015*  *^2^ Subthreshold includes those with sub-threshold BE or BN defined by Micali et al., 2015*  *^3^includes those with full-threshold BED, BN, AN, or PD, and sub-threshold BED and BN, and those with EDNOS and at-risk for EDs, defined by Micali et al., 2015*  *^4^Any age is collapsed across ages 14, 16, or 18. For example, any age full threshold is individuals who had full threshold at 14, 16, and/or 18.* | | | | | | | | | |
